# A variance QTL approach to uncover gene-fish oil supplement interaction loci for 14 circulating unsaturated fatty acid traits

**DOI:** 10.64898/2026.04.13.26350791

**Authors:** Susan Adanna Ihejirika, Eunice Stephen, Kaixiong Ye

## Abstract

Gene-environment interactions (GEI) contribute to circulating polyunsaturated fatty acid (PUFA) and monounsaturated fatty acid (MUFA) profiles. GEI may partly explain differences in trait variance across genotype groups. To identify GEI for circulating unsaturated fatty acids, we adopted a two-stage strategy. First, we detected quantitative trait loci associated with trait variance (vQTLs). Second, we tested these vQTLs for interaction with fish oil supplements (FOS). We performed genome-wide vQTL screens for 14 plasma PUFA and MUFA phenotypes in a UK Biobank subset of 200,478 participants. At the genome-wide significance threshold (*p* < 5.0 x 10^-8^), we identified 172 vQTL-trait pairs across all 14 traits, and 16 of these vQTLs had no marginal genetic effect on the corresponding trait. We found 46 non-overlapping loci across all phenotypes, with an average of 12 vQTLs per trait. Omega-6% and PUFA% had the most independent vQTLs (*N* = 24) while DHA% and Omega-3% had the least (*N* = 1 and 2, respectively). For each of the 172 vQTL-trait pairs, we tested the interaction effect of the vQTL with FOS on the corresponding trait. We found six significant interaction signals in DHA, DHA%, Omega-3, Omega-3%, LA, and Omega-6/Omega-3 ratio around the *FADS1/2, ZPR1*, and *SUGP1/TM6SF2* genes. Our results provide a comprehensive resource of vQTLs and gene-FOS interactions shaping the circulating levels of unsaturated fatty acids.

## Introduction

Unsaturated fatty acids are of varying length and contain at least one carbon double bond.^1^ Monounsaturated fatty acids (MUFAs) contain only one carbon double bond, while polyunsaturated fatty acids (PUFAs) contain two or more. Dietary or circulating PUFAs and MUFAs have been associated with improved outcomes in various diseases, such as cardiovascular disease, diabetes, and obesity.^1,2^ MUFAs can be synthesized endogenously or derived from the diet.^3^ PUFAs fall into two major groups, omega-3 and omega-6 PUFAs, depending on the position of the first carbon double bond from the end of the hydrocarbon chain with the methyl group. Essential PUFAs, including the omega-3 alpha-linolenic acid (ALA) and the omega-6 linoleic acid (LA), can only be derived from diet.^1^ ALA and LA could be endogenously converted by desaturation and elongation enzymatic reactions into long-chain omega-3 and omega-6 fatty acids.^1^ Long-chain omega-3 PUFAs include eicosapentaenoic acid (EPA) and docosapentaenoic acid (DHA), while arachidonic acid (AA) is an example of a long-chain omega-6 PUFA. MUFAs and PUFAs could be obtained from both plant and animal products.^4^ For PUFAs, omega-6 LA accounts for about 85% of all dietary PUFA intake in the modern western diet.^5^

There are ongoing efforts to characterize the genetic architecture of circulating fatty acids.^6–11^ Fully elucidating this genetic architecture requires accounting not only for marginal genetic effects, as evaluated in typical genome-wide association studies (GWAS), but also for gene-environment interactions (GEI). GEI occur when the effect of a genetic variant on a phenotype differs depending on the environmental context.^12^ GEI are non-negligible in understanding the underlying genetic architecture of complex traits and have been recognized as a key mechanism contributing to phenotypic variance in complex human traits.^13^ More specifically for plasma PUFA and MUFA phenotypes, diet has been identified as a key environmental factor modulating genetic effects on these traits.^6,8^ For example, gene-diet interactions in PUFA and MUFA phenotypes are evident at genetic loci involved in converting dietary substrates into circulating forms and downstream bioactive products, such as the *SCD* gene and the *FADS1/FADS2* cluster, which are involved in MUFA and PUFA biosynthesis, respectively.^8,14–16^

In general, the detection of GEI signals has been limited by low statistical power in genome-wide interaction studies (GWIS), the requirement of larger sample sizes than those required to detect marginal genetic effects in GWAS, modest interaction effects, and inaccuracies in exposure measurement.^17–19^ One approach to improve GEI discovery is to capitalize on the genetic effects on the variance of traits. GEI are a likely explanation for the differential phenotypic variance observed across groups when individuals are stratified by genotype at a specific locus.^20^ This proposed approach involves two stages. The first stage is a genome-wide scan to identify loci associated with the variance of the phenotype of interest. These loci are known as variance quantitative trait loci (vQTLs).^20–23^ The second stage is to explicitly test these vQTLs for GEI effects. Uncovering vQTLs informs the search for GEI by prioritizing loci that may harbor relevant GEI, thereby reducing the statistical and computational burden of explicit interaction analysis across the genome.^24^

We aimed to identify genomic loci that shape circulating unsaturated fatty acid profiles through gene-fish oil supplement (FOS) interactions. Though FOS compositions may vary, they are an accessible source of long-chain omega-3 PUFAs, particularly EPA and DHA.^25^ Our analysis builds on our recent large-scale GWIS of FOS in circulating plasma levels of 14 PUFA and MUFA traits.^6^ This study highlighted the *FADS* cluster and the *GPR12* gene. To identify additional interacting loci, we leveraged the UK Biobank cohort to apply the two-stage GEI identification strategy. In stage one, we performed vQTL screens for the same 14 traits. In stage two, for every vQTL of a trait, we explicitly tested whether FOS modifies the vQTL-trait association. Our study expands the scope of investigations into gene-dietary PUFA interaction and contributes to genome-informed personalized nutrition plans that involve unsaturated fatty acids. This study also provides a valuable resource of genetic effects on the variance of PUFA and MUFA traits available in the UK Biobank, which can be further interrogated for underlying GEI of interest.

## METHODS

### Study population

The UK Biobank is a large, prospective cohort of approximately 500,000 United Kingdom-based volunteers recruited between 2006 and 2010, with participants aged 40 to 69 years at the time of recruitment.^26^ For our study, we included up to 200,478 unrelated participants of European ancestry who had no mismatch between genetic and self-reported sex, had no sex chromosome aneuploidy, were not outliers for heterozygosity and missing genotype rate, did not have a high degree of kinship (ten or more third-degree relatives) in the cohort, had a genotype missingness rate of < 5% per individual, and had not withdrawn consent as of February 22^nd^, 2022. The UK Biobank received ethical approval from the National Research Ethics Service Committee North West–Haydock (reference ID 11/NW/0382). The UK Biobank approved this study under Project ID 48818.

### Genotype Data and QC

Information about the genotype data and the initial quality control (QC) process used in this study has been previously described in detail.^27^ We included autosomal variants with an imputation quality score (INFO) > 0.5 and a minor allele frequency (MAF) > 1%. We excluded variants with a genotype missingness rate > 2% per variant or a Hardy-Weinberg equilibrium (HWE) *p*-value < 1.0 x 10^-6^ using PLINK2 alpha-v2.3.^28^ After genotype QC, a total of 7,713,873 SNPs were included in the first stage of our study, the vQTL analysis.

### Phenotype Data and QC

Plasma unsaturated fatty acid levels were measured in a randomly selected subset of about 280,000 participants for the Phase Two release of NMR metabolomic data. Our phenotypes of interest were the absolute circulating levels of total omega-3 fatty acids (referred to as Omega-3), DHA, total omega-6 fatty acids (Omega-6), LA, PUFAs, and MUFAs; their relative percentages in total fatty acids (respectively, referred to as Omega-3%, DHA%, Omega-6%, LA%, PUFAs%, and MUFAs%); and the PUFAs/MUFAs ratio and the Omega-6/Omega-3 ratio.

### Dietary Data

Our study focused on FOS as the dietary exposure of interest. At recruitment, UK Biobank volunteers (*N* = 497,666) completed a dietary survey, indicating how frequently they consumed certain food and drink items. The survey question related to FOS intake was “Do you regularly take any of the following? (You can select more than one answer)”, where “fish oil (including cod liver oil)” was one of the options available for selection (Data Field 6179).

### vQTL analysis

In stage one of our study, we implemented the median-based Levene’s test, as provided in the OSCA software package, to perform vQTL analysis.^22^ The median-based Levene’s test compares absolute deviations from the group medians across genotype groups. We split our dataset into two groups based on sex. For each raw trait value, we adjusted for age and the first 10 genetic principal components and removed outliers by filtering values with a difference from the mean exceeding 5 standard deviations (SDs). Residuals were then standardized to Z-scores in each sex group. For our analysis, we combined the sex-stratified datasets into one after QC and processing.

To interpret the results, we used a genome-wide significance threshold of *p* < 5.0 × 10^-^□. Primary signals were defined using FUMA^29^ with default parameters. We also used FUMA’s GENE2FUNC feature to evaluate tissue specificity and pathway enrichment for candidate genes mapped to independent vQTL loci. Independent lead SNPs were annotated using the WGS Annotator pipeline (version 0.95),^30^ which implemented ANNOVAR^31^ for Ensembl and RefSeq gene annotations, as well as ENCODE-based functional annotations.^32^ For our vQTL and additive QTL overlap and comparisons, we used the publicly available summary statistics of the recent large-scale GWAS of fatty acid traits by Sun et al.^7^ Independent signals were also defined with FUMA. We checked for vQTL and QTL overlap on a trait-by-trait basis, using a window of ±250kb around the lead SNP.

### Colocalization analysis

We performed multi-trait colocalization analysis to identify shared loci and candidate causal variants across our 14 traits of interest, using HyPrColoc (version 1.0).^33^ We used the default priors of *P* = 1 × 10^−4^ for a SNP’s association with a single trait and *P*_c_ = 0.02 as the conditional probability of association with a second trait given a prior association. We defined a significantly colocalized region with a posterior probability (PP) of > 0.7.

### GEI analysis

In our previous large-scale gene-FOS GWIS, we used the Gene-Environment interaction analysis in Millions of samples (GEM) tool^34^ to fit additive generalized linear models and test for interaction effects (1-df test) in 14 rank-based inverse normal-transformed (RINT) PUFA and MUFA phenotypes.^6^ In that study, we adjusted for age, sex, age-by-sex, and the first 10 genetic principal components. For stage two of this study, we extracted vQTL-trait pairs that exhibit significant gene-FOS interactions for the corresponding trait based on the summary statistics of our gene-FOS GWIS. To determine the significance of interactions in each trait, we applied different Bonferroni-corrected significance thresholds, estimated from *p* < 0.05 adjusted for the number of independent vQTLs tested in the trait of interest. The same method was used to determine the significance thresholds when testing QTLs for interaction effects in each trait.

## Results

### Cohort demographics

Table S1 summarizes the characteristics of participants who met all genotype, phenotype, and dietary QC requirements (see Methods and Figure S1). Of the *N* = 200,060 participants included, approximately 31.8% (*N* = 63,711) took fish oil regularly. Females comprised 53.5% of our cohort, and the mean age at recruitment was 56.8 ± 8.0 (SD) years.

### Mapping vQTLs for circulating PUFAs and MUFAs

The first stage of our study involved performing genome-wide vQTL scans in 14 PUFA and MUFA traits using Levene’s median-based test (Figure S2). With LD-based clumping and merging of overlapping loci within a ±250kb window, we identified 172 independent significant loci at the genome-wide significance threshold of *p* < 5 × 10^-8^ (Figure 1A, Tables S2 and S3) across all traits. Omega-6% and PUFAs% yielded the largest number of independent vQTLs (*N* = 24) while DHA% and Omega-3% had the least (*N* = 1 and 2, respectively). The majority of vQTLs were located within intronic (52.9%) and intergenic (29.8%) regions, consistent with previous findings that trait-associated variants are primarily found in noncoding regions, supporting the hypothesis that gene regulation plays a key role in mediating the effects of vQTLs (Figure S3A). We then annotated the intergenic vQTLs using ENCODE and found that most fell within promoter or promoter flanking regions (Table S4).

**Figure 1.**
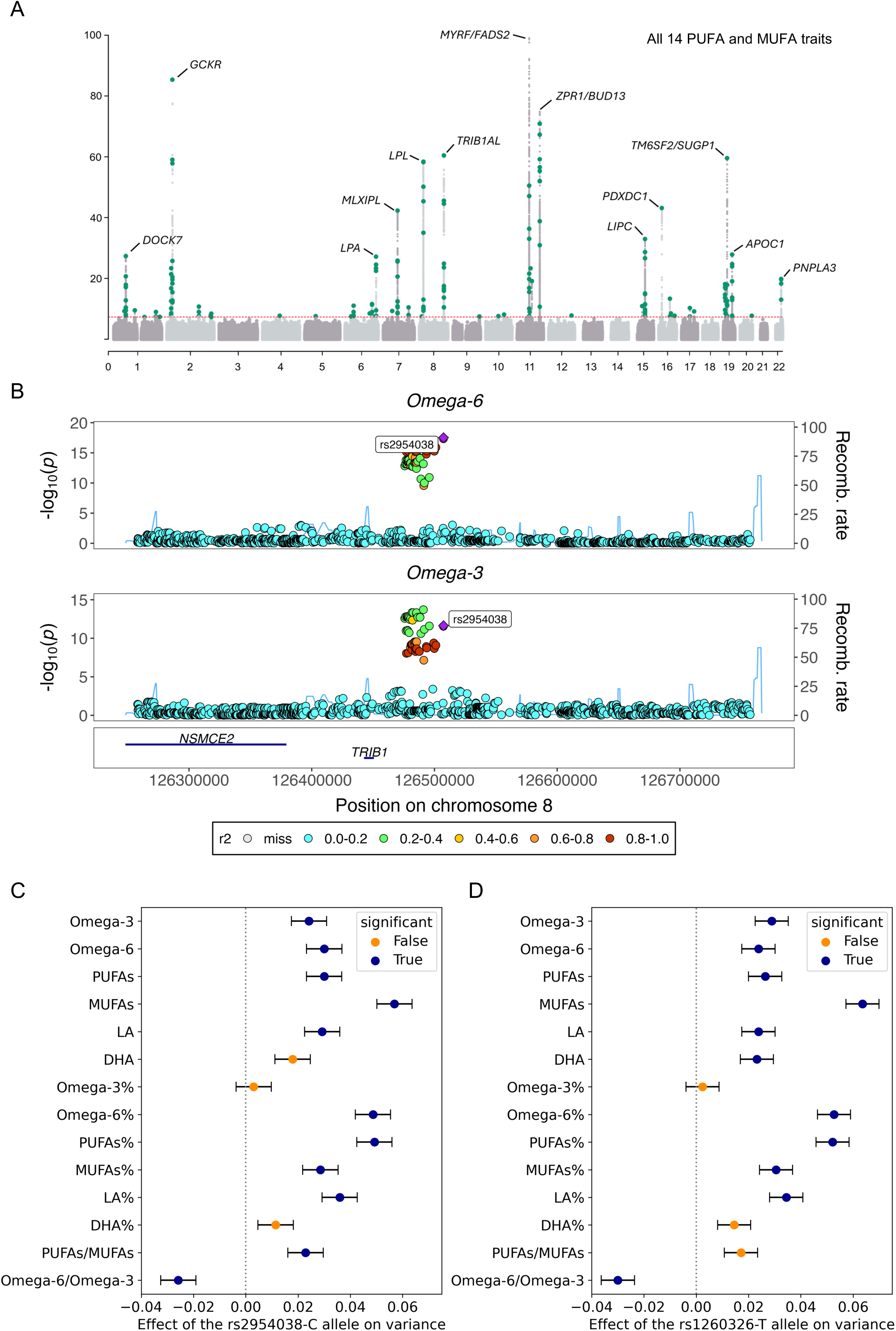
vQTLs for 14 circulating PUFA and MUFA traits. A) Manhattan plot showing 172 genome-wide significant vQTL lead variants for 14 PUFA and MUFA traits. Top SNPs in each region are labeled with the nearest gene. A maximum of −log_10_*P* = 100 was used for visualization purposes. B) Stacked regional Manhattan plots for Omega-3 and Omega-6 showing the most pleiotropic locus in this study and highlighting the top SNP rs2954038. C) The *TRIB1AL* region has significant effects on the variance of 12 out of 14 PUFA and MUFA traits, except omega-3% and DHA%. The rs2954038 SNP was a vQTL in 11 out of 14 traits. D) *GCKR*-rs1260326 has significant effects on the variance of 11 out of 14 PUFA and MUFA traits, except Omega-3%, DHA%, and the PUFAs/MUFAs ratio.

To account for redundancy among overlapping vQTLs, we used a ±250kb window to extract non-overlapping, unique loci across all traits. Of the 46 identified non-overlapping loci, 50% (23 out of 46 loci) were specific to a single trait, indicating trait-specific genetic effects on phenotypic variance. The remaining 50% displayed association with variance in at least two traits (Figure S3B, Table S5). The locus encompassing the SNPs rs2954038, rs2954021, rs2954029, rs2980860, and rs2980888, all mapped to the *TRIB1*-associated lncRNA (*TRIB1AL*) gene, showed the broadest impact, influencing phenotypic variance in 12 of the 14 traits, except for Omega-3% and DHA%. This region was represented by the rs2954038 SNP (chr 8:126507389; C>A; MAF = 0.30), which showed the strongest statistical significance across variants in this region (*p* = 3.29 x 10^-61^ in MUFAs). The rs2954038 SNP itself had significant effects on the variances of 11 traits, except for Omega-3%, DHA, and DHA% (*p* = 0.38, 2.0 x 10^-7^, and 9.3 x 10^-4^, respectively). The rs2954038-C allele had the strongest effect on MUFAs, increasing its variance by 0.06 (se = 0.003) per allele (Figure 1B and 1C). The rs2954038-C allele was associated with a decrease in variance only in the Omega-6/Omega-3 ratio (β = −0.03, se = 0.003). The rs1260326 SNP (chr 12:27730940; T>C, MAF = 0.39), a missense variant mapped to the *GCKR* gene, was a vQTL in 11 traits, except Omega-3%, DHA%, and the PUFAs/MUFAs ratio (*p* = 0.45, 7.32 x 10^-6^, and 1.18 x 10^-7^, respectively). The rs1260326 variant has been associated with a diverse range of traits, including metabolite levels, totaling over 100 human traits and disease associations.^35^ Consequently, the *GCKR* gene is one of the highly pleiotropic loci, not only for marginal genetic effects but also for vQTLs (Figure 1D).

Among the 46 non-overlapping loci, 5 were unique for the Omega-6/Omega-3 ratio, and 1 was unique for the PUFAs/MUFAs ratio. The remaining 17 unique loci are partitioned among MUFAs (N = 5), Omega-6% (N = 4), PUFAs% (N = 3), LA (N = 2), Omega-3 (N = 1), DHA (N = 1), and MUFAs% (N = 1) (Table S5). Categorizing the non-ratio traits into four groups, the group of MUFAs and MUFAs% has a total of 23 vQTLs, the group of PUFAs and PUFAs% has 25, the group of four omega-6 traits has 28, and the group of four omega-3 traits has 12. A total of 10 loci overlapped across these four groups (Figure 2A). Among the 23 loci that were shared by at least two traits, the combination of PUFAs% and Omega-6% shared the most loci (N = 4), while most specific trait combinations (N = 14) shared only one locus. The trait with the most shared loci was PUFAs% (N = 21), while Omega-6% was ranked second (N = 20) and MUFAs third (N = 16) (Figure 2B).

**Figure 2.**
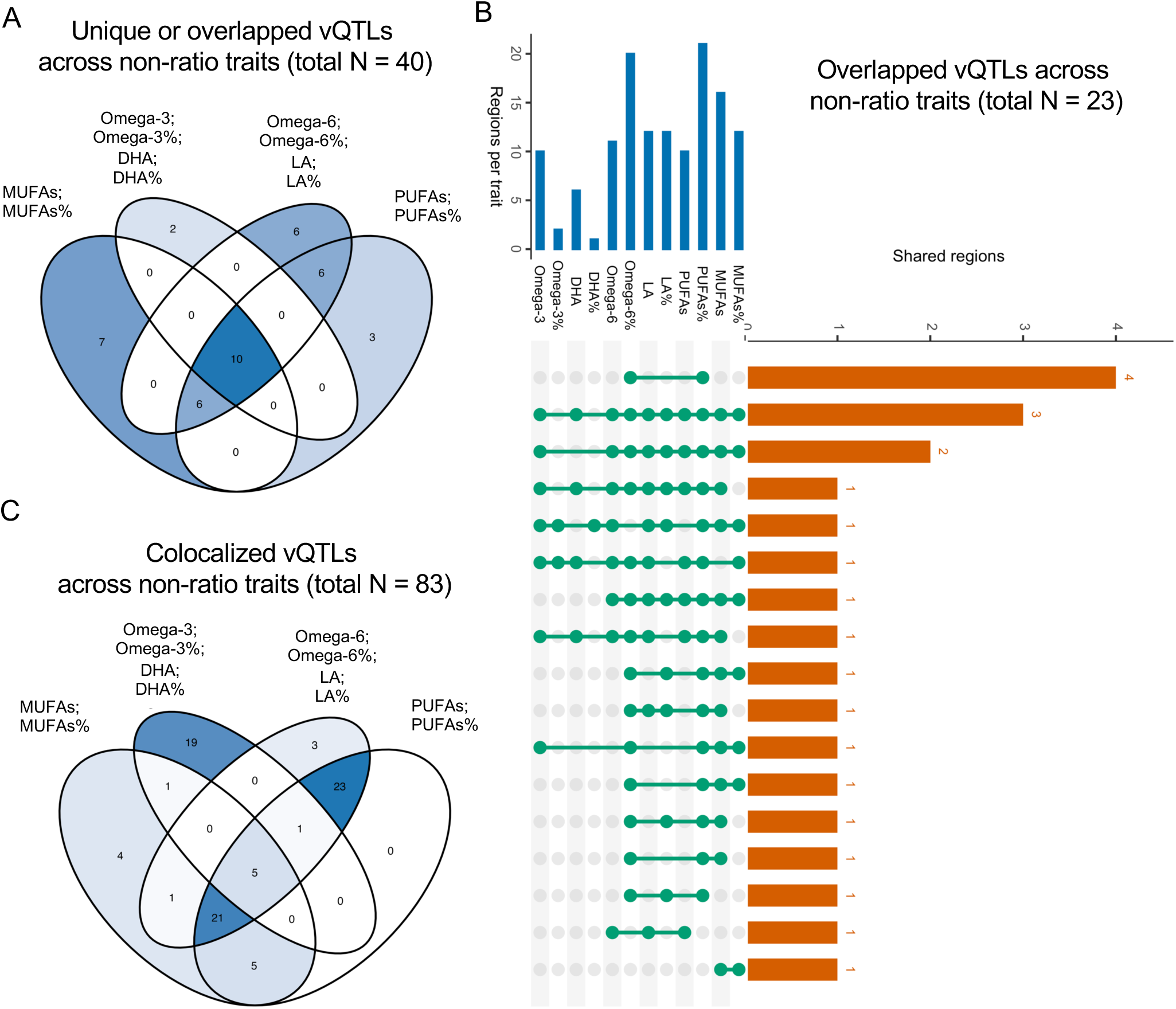
Overlapped or colocalized vQTLs across traits. A) A Venn diagram for 40 nonoverlapping loci across 12 non-ratio traits. The six loci specific to the ratios of Omega-6/Omega-3 or PUFAs/MUFAs were excluded from this visualization. B) An UpSet plot for 23 overlapped vQTLs across 12 non-ratio traits. C) A Venn diagram for 83 colocalized regions across 12 non-ratio traits.

To systematically investigate shared vQTLs across all traits while accounting for linkage disequilibrium (LD), we conducted multi-trait colocalization analysis to identify loci harboring causal variants that may influence phenotypic variance in multiple traits. We observed 83 significant clusters, each with a corresponding candidate causal variant, where two or more traits colocalized at PP > 0.7 (Figure S4, Table S6). The rs1260326 *GCKR* locus was shared across the largest cluster of traits, consisting of 13 out of 14 traits. Categorizing the non-ratio traits into four groups, the group of MUFAs and MUFAs% was involved in 37 colocalized loci, but with only 4 unique to this group. The group of PUFAs and PUFAs% was involved in 55 colocalized loci, but all are shared with at least one of the other three groups. A total of 19 loci were colocalized in the group of four omega-3 traits, while 3 loci were in the group of four omega-6 traits. A total of 5 loci were colocalized for all four groups of traits (Figure 2C). To list some examples, the GDP-Mannose 4,6-Dehydratase (*GMDS*) gene, located on chromosome 6 and involved in the synthesis of GDP-fucose and the fucosylation of many proteins^36^, was a shared locus in LA and Omega-6. A colocalized locus for DHA and Omega-3 was the Alpha-Actinin-2 (*ACTN2*) gene, located on chromosome 1, and encodes the alpha-actinin-2 structural protein expressed in skeletal and cardiac muscles.^37^

### vQTLs substantially overlap with additive QTLs for plasma unsaturated fatty acids

The majority of our vQTLs overlapped with additive effect loci (also referred to as QTL loci) previously reported in a large-scale GWAS of PUFA and MUFA traits by Sun et al.^7^ We checked for overlap, on a trait-by-trait basis, between 172 vQTL loci and 1,164 QTL loci (Figure 3A). Of the 172 lead vQTLs across all traits, 90.7% (156 of 172) also had a marginal effect, while only 13.4% of QTL loci contained vQTLs (Table S7). Comparison of effect sizes of the 156 overlapped loci between vQTL and additive QTL loci showed a moderate positive relationship (r = 0.53), suggesting that variants with stronger effects on mean trait levels also tend to have stronger effects on trait variance levels (Figure 3B). This pattern aligns with previous findings that loci with interaction effects in a phenotype of interest tend to cluster in regions with known functional connections to the phenotype.^21,24,38^ The region surrounding the rs7943728 vQTL showed both the most significant vQTL effect (*p*_vqtl_ = 1.7 x 10^-322^) and QTL effect (*p*_qtl_ = 5 x 10^-324^), both in the Omega-6/Omega-3 ratio. From the GTEx eQTL dashboard, we found that rs7943728 is an eQTL for *FADS2*.

**Figure 3.**
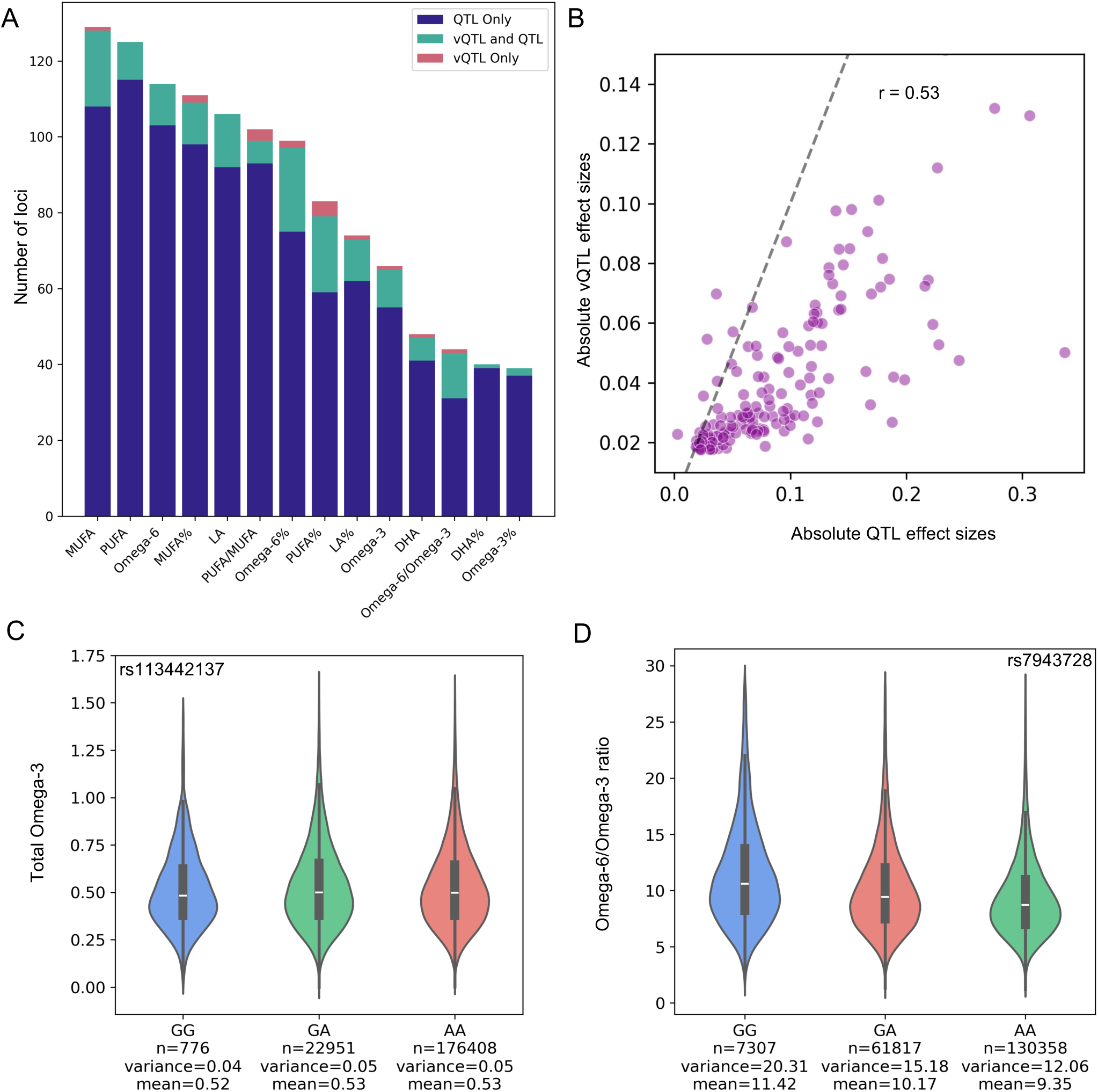
The overlap between vQTL and QTL. A) The counts of significant vQTL and additive QTL for each phenotype. Blue indicates QTL-only loci, green indicates vQTL and QTL shared loci, and red indicates vQTL-only loci. B) Comparison of vQTL and QTL effect sizes for loci with both effects in at least one of the 14 traits. The dashed line represents y = x. C) An example of a vQTL-only locus (rs113442137) for total Omega-3 levels. D) An example of a vQTL and QTL shared locus (rs7943728) for the Omega-6/Omega-3 ratio. The center white line of the violin plot shows the mean trait level.

There was a total of 16 vQTL-only loci across DHA (*N* = 1), Omega-3 (*N* = 1), Omega-6% (*N* = 2), LA% (*N* = 1), MUFAs (*N* = 1), MUFAs% (*N* = 2), PUFAs% (*N* = 4), and the PUFAs/MUFAs (*N* = 3) and Omega-6/Omega-3 (*N* = 1) ratios (Table S8). Omega-3%, DHA%, LA, Omega-6, and PUFAs had no vQTL-only loci. PUFAs% had the highest number of vQTL-only sites, with 4 of its 24 vQTLs lacking QTL effects. Figures 3C and 3D show two examples of vQTL loci with and without additive effects on trait mean in Omega-3 and the Omega-6/Omega-3 ratio, respectively.

### Tissue-specific expression of candidate genes in vQTLs

We used the GENE2FUNC module in FUMA to assess the enrichment of differential gene expression across tissues for candidate genes mapped to vQTLs. This analysis was performed separately for each trait (Figure S5, Table S9). We found significant tissue-specific expression of our vQTL-mapped genes in the brain, esophagus, liver, adrenal gland, vagina, whole blood, colon, and EBV-transformed lymphocytes, with varying patterns of upregulation and downregulation in DHA%, LA%, Omega-6%, and PUFAs%. In the context of vQTLs, tissue-specific expression suggests that these tissues may be of interest when looking for context-specific expression. We also conducted gene set enrichment analyses across all 14 traits and identified enriched pathways for hallmark gene sets, except for the absolute concentrations of DHA and PUFAs. Cholesterol homeostasis and MTORC1 signaling were generally shared enriched pathways across both omega-3- and omega-6-related traits. Gene sets associated with omega-3-related traits were also enriched in bile acid metabolism, while heme metabolism was enriched for gene sets in omega-6-related traits (Table S10).

### Tests for gene-FOS interactions reveal trait-specific FOS-interacting loci

In the second stage of our study, we tested the 172 lead vQTL-trait pairs for interaction with FOS in the corresponding PUFA and MUFA traits, defining significance at trait-specific thresholds (Table S11). We observed six Bonferroni-corrected, significant GEI in DHA, DHA%, Omega-3, Omega-3%, the Omega-6/Omega-3 ratio, and LA, respectively (Table S12). The five significant variants in omega-3-related traits were mapped to the *FADS/MYRF* region and the *ZPR1* gene, while the one variant in LA was mapped to the *SUGP1/TM6SF2* region. We found that the G allele of the rs964184 SNP (chr 11:116648917; G>C, MAF = 0.14), in the 3’-UTR of the *ZPR1* gene, was associated with increased DHA, having a larger effect in individuals taking FOS (β = 0.05, se = 0.008) than in those who do not use FOS (β = 0.02, se = 0.005) (Figure 4A). From genotype stratification, we found that FOS had varying effects across the three genotype groups: β = 0.57, 0.47, and 0.46 SD units; se = 0.037, 0.001, and 0.005, respectively, for G/G, G/C, and C/C (Figure 4B).

**Figure 4.**
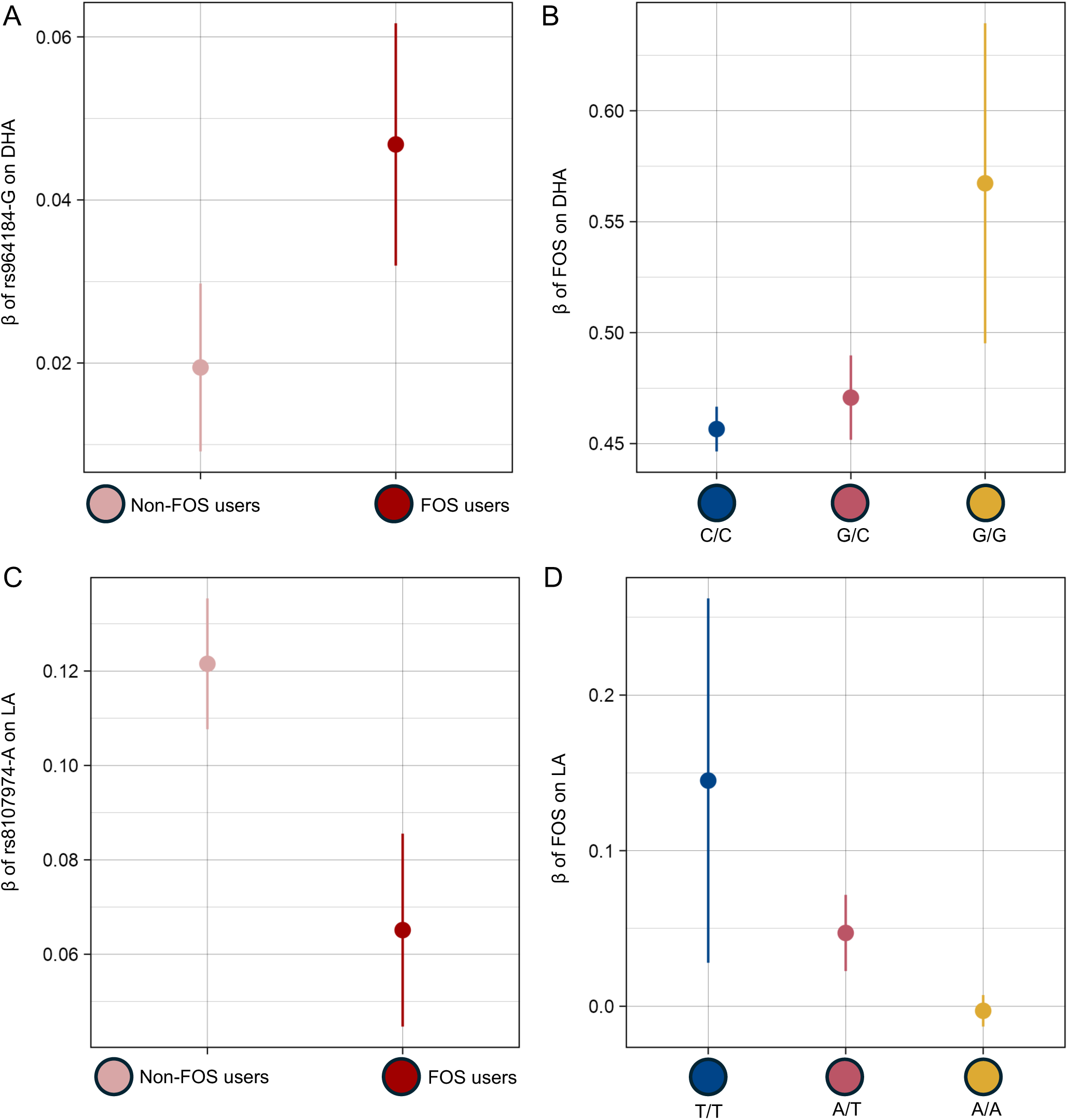
Stratified analysis for two significant FOS-interacting loci. A) Marginal genetic effect of rs964184 on Omega-3 in FOS users and non-users. B) The effect of FOS on total DHA across rs964184 genotype groups. C) Marginal genetic effect of rs8107974 on LA in FOS users and non-users. D) The effect of FOS on total LA across rs8107974 genotype groups. All error bars represent 95% confidence intervals.

The rs35473591-C variant (chr 11:61586328; C>CT, MAF = 0.34), which is the top SNP in the *FADS/MYRF* region, had a smaller effect on Omega-3 in FOS users (β = 0.30, se = 0.005) than in those who did not use FOS (β = 0.36, se = 0.004) (Figure S6A). Across the C/C, C/CT, and CT/CT genotype groups, FOS had effects of β = 0.39, 0.43, and 0.50, se = 0.007, 0.007, and 0.014, respectively (Figure S6B). We previously reported the modifying effect of FOS on the relationship between rs35473591 and Omega-3%.^6^ Lastly, the A allele of the *SUGP1/TM6SF2-*rs8107974 variant showed a stronger effect on LA in non-users (β = 0.12, se = 0.007) than in FOS users (β = 0.07, se = 0.01) (Figure 4C). From genotype-stratified analysis at this locus, we found that FOS had no effect on LA in A/A homozygotes but was positively associated with LA in the T/T and A/T groups (β = 0.15 and 0.05, se = 0.06 and 0.01, respectively) (Figure 4D). Overall, with multiple testing corrections, significant interactions with FOS were identified for 3.49% (6 of 172) of the trait-vQTL pairs tested. At the nominal significance of *p* < 0.05, we identified 24 suggestive interaction loci (Table S13).

We then tested the 1,164 previously identified lead QTLs for gene-FOS interactions at trait-specific thresholds (Table S11). We found five significant GEIs across three unique variants and five traits: LA, PUFAs, Omega-3%, the Omega-6/Omega-3 ratio, and Omega-6 (Table S14). These variants were also mapped to the *FADS1/FADS2* gene cluster and the *SUGP1/TM6SF2* region. These interactions account for 0.43% (5 of 1,164) of the QTL-trait pairs tested. The proportion of vQTLs that had an interaction effect was 8.12-fold higher than the proportion of additive QTLs. Lastly, using a one-sided binomial test across all 14 traits, we observed significant enrichment of gene-FOS interaction signals among vQTLs, with 24 out of 172 nominally significant loci (14%; *p* = 5.8 × 10^-6^) showing GEI with FOS, substantially higher than the null expectation under *p* = 0.05. These findings support the usage of vQTLs as prioritized candidate loci for interaction analysis.

### A closer look at *KSR2-*rs7954783, a vQTL without QTL effect

Focusing on the vQTL loci without QTL effects, the rs7954783 locus (chr 12:118344424; C>A, MAF = 0.087) was the only vQTL out of 16 vQTL-only sites to reach nominal significance for interaction at a threshold of *p* < 0.05 (Table S13). An explanation for why a locus may have a vQTL effect but not a QTL effect is that it is a ‘directionally discordant’ GEI.^24,39^ Directionally discordant GEI are defined as vQTLs that may lack additive marginal effects because they show opposing effects in stratified groups of an environmental exposure of interest.^24,39^ The rs7954783 SNP is located in an intron of the *KSR2* (kinase suppressor of Ras2) gene, which is known to regulate the Ras/MAPK signaling pathway and energy homeostasis and has been associated with metabolic disorders, including obesity.^18,40^ The rs7954783-C allele had a significant vQTL effect (*p* = 1.55 × 10^-8^), associated with decreased variance in the Omega-6/Omega-3 ratio (β = −0.07, se = 0.01; Figure 5A) and interacting significantly with FOS (*p* = 0.01) to influence this ratio (Table S13). From stratified genetic association analysis by FOS intake, we observed that at *p* < 0.05, the rs7954783-C allele lacked an effect on the mean ratio in FOS users (*p* = 0.6) but showed a significant effect in non-FOS users (*p* = 0.03, β = −0.041, se = 0.01; Figure 5B). The FOS had different effects on the Omega-6/Omega-3 ratio in C/C homozygotes (β = −0.47, se = 0.005), C/A heterozygotes (β = −0.50, se = 0.03), and A/A homozygotes (β = −1.13, se = 0.30) (Figure 5C).

**Figure 5.**
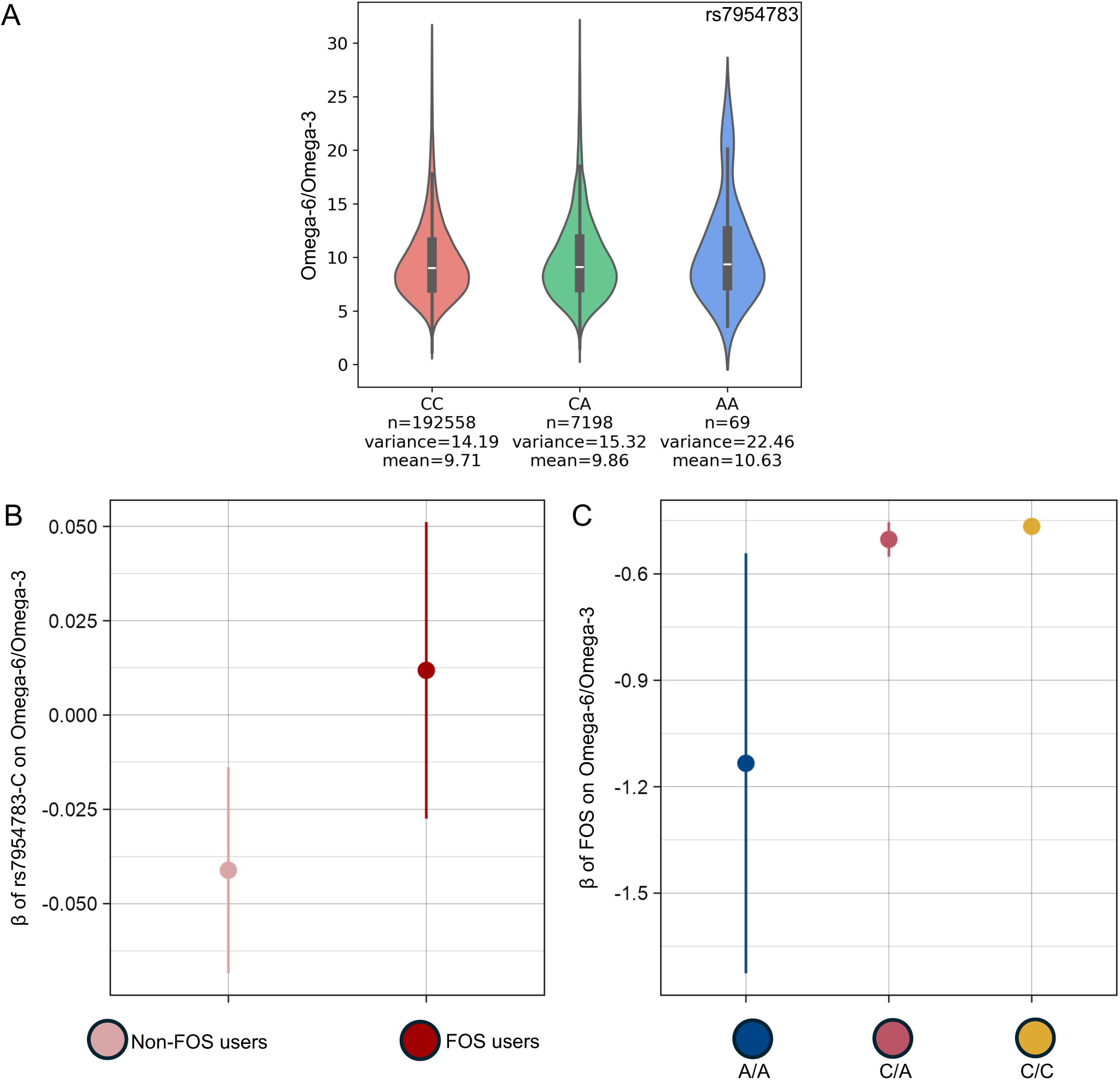
A vQTL-only locus. A) The rs7954783 SNP is a vQTL with no QTL effect on the corresponding trait. The center white line in the violin plot shows the mean Omega-6/Omega-3 ratio. B) Marginal genetic effect of rs7954783 on the Omega-6/Omega-3 ratio in FOS users and non-users. C) The effect of FOS on the Omega-6/Omega-3 ratio across rs7954783 genotype groups. All error bars represent 95% confidence intervals.

To further investigate how the rs7954783-C allele influences the Omega-6/Omega-3 ratio, we explored its estimated effects on Omega-3 and Omega-6 from previous GWAS summary statistics. We found that at *p* < 0.05, the rs7954783-C allele is positively associated with Omega-3 (*p* = 0.01, β = 0.03, se = 0.013) but not with Omega-6 (*p* = 0.11, β = 0.02, se = 0.012).^7^ In summary, we did not observe opposing effects at the rs7954783 locus across the two exposure strata. This observation suggests an alternative explanation for loci that show vQTL effects but no detectable QTL effects on the same phenotype. In this case, the rs7954783-C allele may exert a QTL effect on the mean Omega-6/Omega-3 ratio in non-FOS users, but not in FOS users, leading to attenuation of the effect on the trait mean when the two exposure groups are combined. Overall, these results highlight that stratifying analysis by FOS usage revealed group-specific genetic effects not detected in typical GWAS and suggest a modulating effect of the rs7954783 genotype on the association between FOS and the Omega-6/Omega-3 ratio.

## Discussion

PUFAs and MUFAs are integral components of the human diet and are crucial for human health. It is important to investigate how our PUFA intake and genetic makeup contribute to diverse circulating levels of unsaturated fatty acids among individuals. In this study, we applied a two-stage approach and leveraged a cohort of UK Biobank participants to characterize gene-FOS interacting loci that influence 14 plasma PUFA and MUFA traits. We first identified vQTLs associated with the variance of each trait and tested them for interaction with FOS intake in their corresponding traits. Through this two-step strategy, we identified 172 independent vQTLs (46 non-overlapping) across all 14 traits. We also identified shared, putative causal variants across traits and tested for tissue-specific expression and pathway enrichment of candidate genes at vQTL loci. In the second stage of our study, we uncovered six significant gene-FOS interactions in DHA, DHA% Omega-3, Omega-3%, LA, and the Omega-6/Omega-3 ratio at three independent loci, including the *MYRF/FADS1/FADS2*, *ZPR1* (near the *APOA5* gene cluster), and *SUGP1/TM6SF2* loci. In our previous gene-FOS GWIS, we identified interactions at the *FADS1/FADS2* cluster in DHA, DHA%, Omega-3, Omega-3%, and the Omega-6/Omega-3 ratio, and highlighted the differential effect of the top SNP, rs35473591, on Omega-3% in the C/C, C/CT, and CT/CT genotype groups.^6^

We found that 90.7% of our lead vQTL loci also harbored an additive genetic effect in their associated trait and had been identified by previous GWAS.^7^ This substantial overlap suggests that prioritizing loci with marginal genetic effect can contribute to GEI testing.^41^ However, this approach may overlook SNPs with very small interaction effects. The Levene’s median test is powered to prioritize loci with small interaction effects.^20^ Our results provide further evidence that such variance heterogeneity screens are effective, as we observed an 8.12-fold enrichment of GEI signals when screening vQTLs compared to additive QTLs. This is consistent with recent large-scale analyses, which found a 10- and 5-fold enrichment of GEI signals in vQTLs compared to QTLs.^24,39^ We identified 16 loci with effects on only trait variance. Of the 16 vQTL-only sites, none of the vQTL-trait pairs reached Bonferroni-corrected statistical significance for gene-FOS interactions in their associated phenotype. Future analyses could test these vQTLs for GEI effect with other relevant exposures, such as other dietary sources of PUFAs and MUFAs. Overall, taking a vQTL prioritization approach enabled the discovery of two additional gene-FOS interactions that were not uncovered in the previous GWIS.^6^

Pathway analysis of vQTL-mapped genes highlighted shared programs across traits, e.g., cholesterol homeostasis and mTORC1 signaling. mTORC1 is a central driver of hepatic lipogenesis via activation of SREBP-1c. In human and model systems, DHA can suppress mTORC1/SREBP-1c signaling, whereas AA and LA activate mTORC1.^42–45^ There were also group-specific signals, with omega-3-related traits being enriched for bile acid metabolism, while omega-6-related traits showed heme metabolism signatures, consistent with known roles in lipid handling and inflammatory biology. We emphasize that these results reflect bulk-tissue expression and gene-set enrichment, rather than causal assignments. It has been established that most candidate genes at GWAS loci have tissue-specific expression patterns;^46^ hence, we hypothesized that candidate genes at vQTL loci follow a similar pattern. Our tissue enrichment analysis of vQTL-mapped genes revealed significant enrichment in metabolically and functionally relevant tissues, most notably the liver, brain, whole blood, and lymphoid cells, as well as select gastrointestinal and endocrine tissues. Enrichment patterns varied by PUFA phenotype, supporting biological specificity. For example, DHA%-associated vQTL genes were upregulated in the brain’s putamen and hippocampus. DHA’s presence and function have been well-documented in the brain.^47^

Our GEI analysis highlighted six significant interactions, mapped to three independent loci. The *MYRF* rs509360 and rs7943728 SNPs are in LD with variants in the *FADS1/2* cluster. The *FADS1-FADS2* cluster is a well-established locus in fatty acid biosynthesis, as it converts dietary precursors to long-chain PUFAs. This cluster encodes the delta-5 and delta-6 desaturase enzymes that introduce cis double bonds at the delta-5 and delta-6 positions, respectively, in PUFAs.^48^ Genetic variants in *FADS1/2* affect endogenous synthesis of long-chain PUFAs.^7,9,10^ These variants have been associated with varying conversion efficiency of 18-carbon precursors (LA, ALA) to long-chain PUFAs (AA, EPA, DHA). Our results and prior studies^6,8^ indicate that these genetic effects can be modulated by omega-3 intake. Similarly, we observed that the examined alleles of the *FADS/MYRF* variants (e.g., rs35473591-C) were associated with smaller increases in Omega-3 among FOS users, consistent with the hypothesis that fish oil provides an exogenous source of these fatty acids, diminishing the need for endogenous synthesis and thus the effects of genetic variants in this pathway.

We observed another significant interaction between the *ZPR1*-rs964184 variant and FOS on DHA. The *ZPR1* gene encodes a zinc finger protein that is a key player in cell growth and proliferation.^49,50^ The rs964184 SNP is in high LD with variants in the *APOA5-A4-C3-A1* apolipoprotein gene cluster, which is involved in lipid metabolism and triglyceride regulation.^51^ The rs964184 variant has also been repeatedly associated with PUFA and MUFA traits. Our analysis showed that the rs964184-G allele was associated with a greater increase in DHA in individuals taking FOS compared to non-users. The effect size of FOS on DHA increased with each copy of the G allele. In other words, carriers of the rs964184-G allele may derive more benefit from FOS intake in terms of increasing DHA, with G/G homozygotes seeing the most benefit and C/C homozygotes getting the least. The mechanisms underlying the rs964184-FOS interaction have not been elucidated. We hypothesize that genetic differences in relevant lipid pathways affect the efficiency with which dietary omega-3 fatty acids are incorporated into plasma lipids.^52^ Future studies are needed to test this hypothesis.

At 19p13.11, our lead variant, *SUGP1*-rs8107974, lies within the *SUGP1-TM6SF2* LD block. While recent evidence has shown that *SUGP1* variants may be functionally relevant in lipid metabolism,^53^ we interpreted this interaction in terms of the *TM6SF2* locus because *SUGP1*-rs8107974 is correlated with rs58542926, the well-documented *TM6SF2* E167K variant (r^2^ = 1.0).^54^ *TM6SF2* (Transmembrane 6 Superfamily 2) encodes a protein involved in hepatic lipid secretion, and the *TM6SF2* E167K variant reduces very large density lipoprotein (VLDL) export from the liver, influencing blood lipid levels and predisposition to non-alcoholic fatty acid disease (NAFLD).^55^ We observed that the rs8107974-A allele was associated with a smaller increase in LA in FOS users than in non-FOS users.

The GEI effect in the Omega-6/Omega-3 ratio explains why the *KSR2*-rs7954783 variant affects only the variance of the ratio, not its mean. Only with stratification by FOS intake did the underlying QTL effect emerge. The rs7954783-C allele showed a reducing effect on the ratio in non-FOS users but had no significant effect on the ratio in FOS users. One speculative explanation for the observed effect is that, in the absence of exogenous omega-3, the rs7954783-C allele promotes metabolic processes that favor a lower Omega-6/Omega-3 ratio, possibly by enhancing endogenous omega-3 production or retention. The rs7954783-C allele has a significant effect on omega-3 levels, but not on omega-6 levels, supporting this explanation. The precise underlying biological mechanism remains to be elucidated. In addition, this result supports the two-step vQTL prioritization approach for identifying context-dependent SNP effects.

Key strengths of our study include the large sample size and prioritization of SNPs for interaction testing, both of which enhanced power to detect more gene-FOS interactions in PUFA and MUFA traits compared to our previous analysis, which utilized direct genome-wide interaction tests.^6^ This study has some limitations. Firstly, we included only participants of genetic European ancestry because of the limited sample sizes for other ancestries in the UK Biobank. Secondly, our FOS intake data did not include other important quantitative measurements, such as frequency and supplement composition. Lastly, we did not adjust for environmental covariates that may be associated with FOS (e.g., other dietary sources of unsaturated fatty acids and socioeconomic status); thus, the effect estimates may be affected by uncontrolled confounding. In the case of omega-3 PUFAs, however, our previous gene-FOS interaction analysis showed that, even after accounting for oily fish intake, SNP effect estimates remained consistent with those obtained when only FOS were considered.^6^

## Conclusion

Our study uncovered 172 unique genome-wide significant vQTLs for 14 circulating PUFA and MUFA traits. Testing these vQTLs for interactions with FOS, we identified two additional loci that significantly modify the effects of FOS on circulating PUFAs. Our results are publicly accessible and serve as a resource for understanding the effects of SNPs on the phenotypic variance of plasma PUFA and MUFA traits. Prioritized SNPs can be tested for GEI effects with other environmental exposures in future analyses. Our GEI findings can contribute to the development of FOS-aware interaction polygenic scores (iPGS) for these 14 PUFA and MUFA traits, which can be used to stratify populations and inform dietary recommendations. In addition, our vQTL results can contribute to emerging efforts to develop variance polygenic scores (vPGS).^56^ Variance PGS is an approach that incorporates individual differences and can be used in conjunction with PGS to enhance trait and disease risk prediction and stratification.^56^

## Description of Supplemental Information

Supplemental information includes six figures and 14 tables.

## Acknowledgments

This research has been conducted using the UK Biobank Resource under Application Number 48818. We express our heartfelt gratitude to the volunteers and staff who made the UK Biobank dataset possible. Research reported in this publication was supported by the National Institute of General Medical Sciences of the National Institute of Health under the award number R35GM143060 (KY). The content is solely the authors’ responsibility and does not necessarily represent the official views of the National Institutes of Health.

## Data and Code Availability

The 14 sets of vQTL summary statistics will be made available in the GWAS Catalog (https://www.ebi.ac.uk/gwas/). Our previous gene-FOS GWIS summary statistics are available in the GWAS Catalog under accession IDs from GCST90565570 to GCST90565583. Scripts used in this analysis are available at https://github.com/adannasusan/PUFA-vQTL.

