## Supplementary Figures for "A variance QTL approach to uncover gene-fish oil supplement interaction loci for 14 circulating unsaturated fatty acid traits"

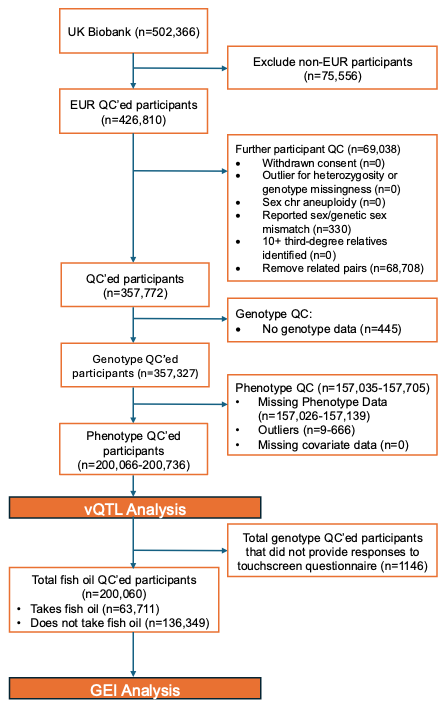

Figure S1. Flowchart of participant inclusion criteria. Individuals included in the GEI analysis were adapted from our previous FOS GWIS.^1^

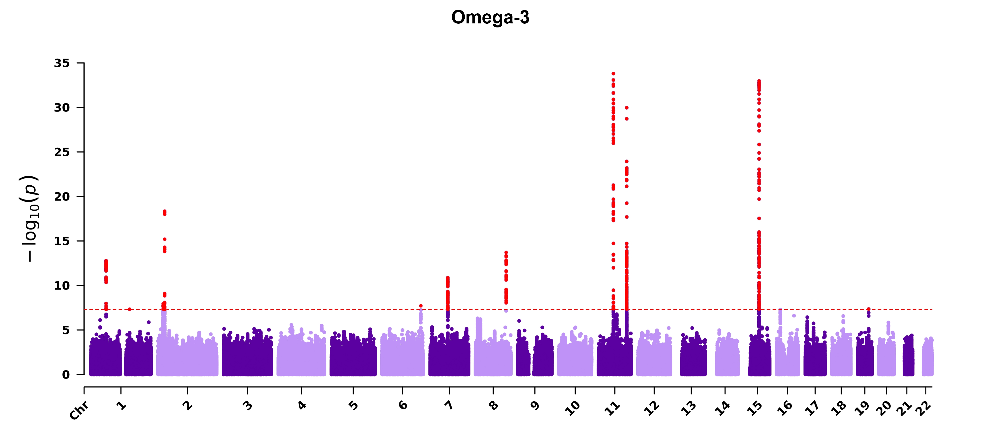

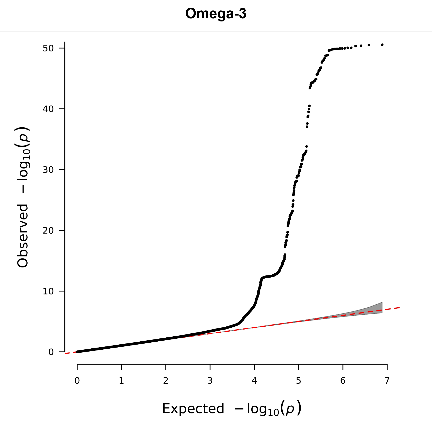

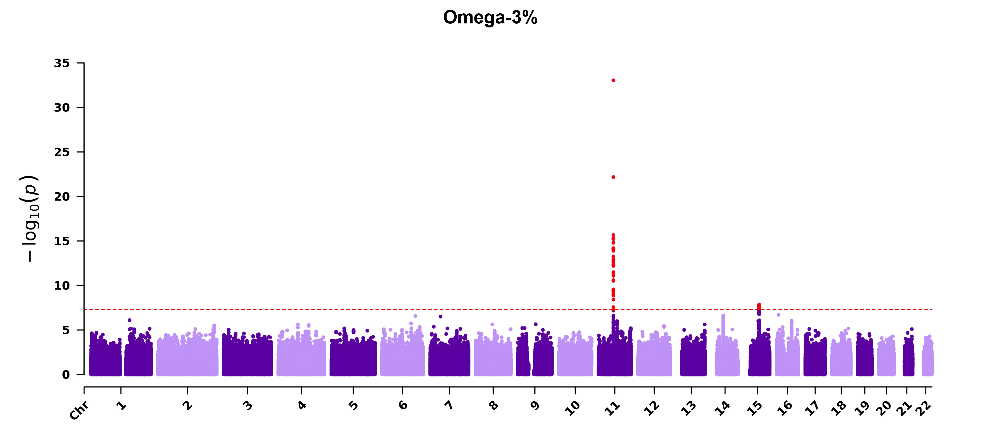

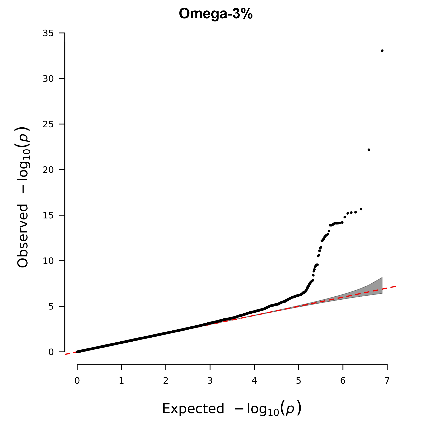

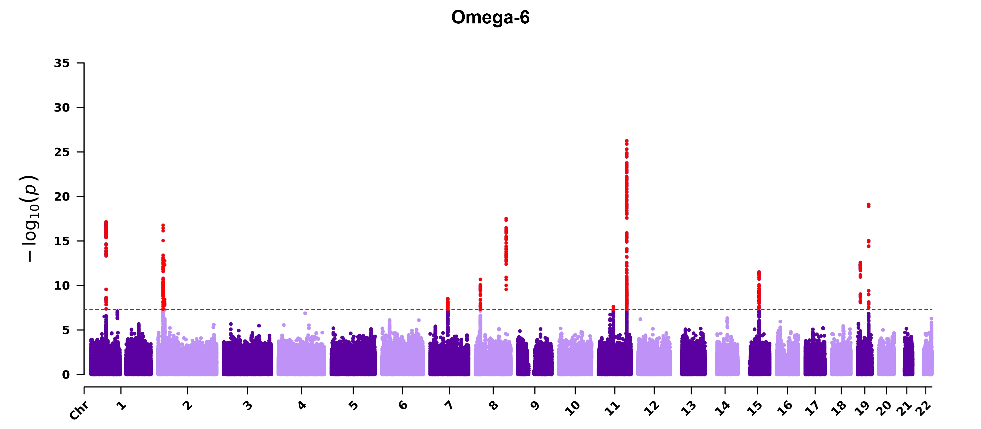

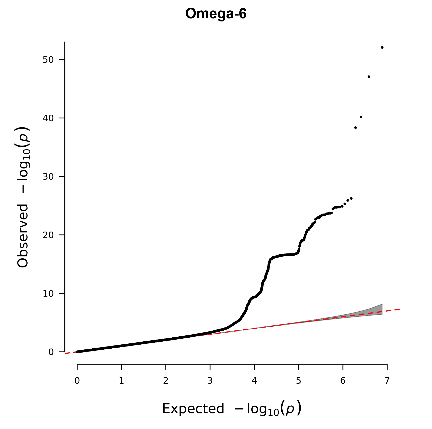

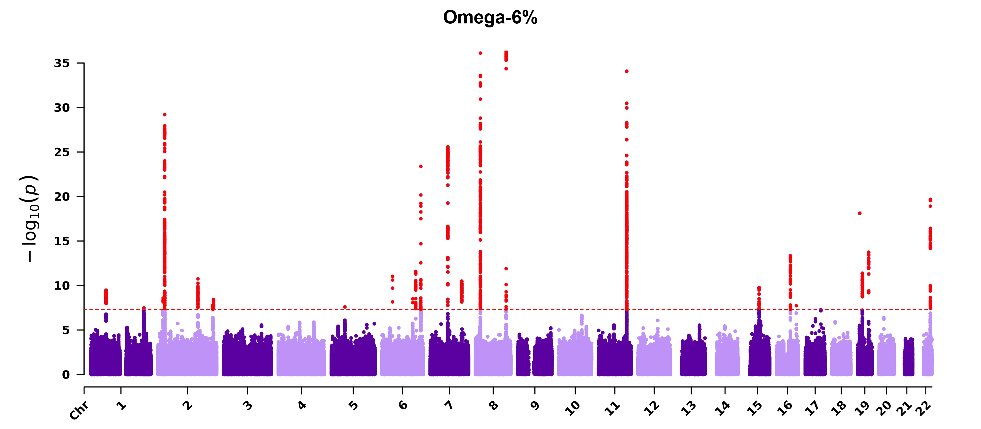

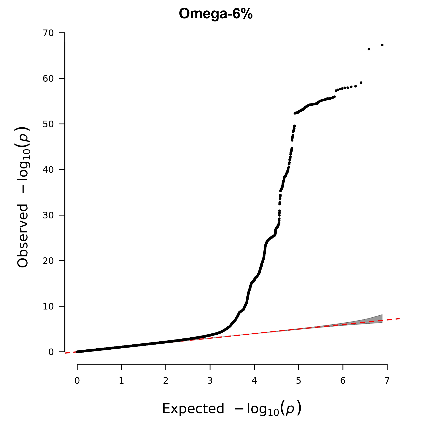

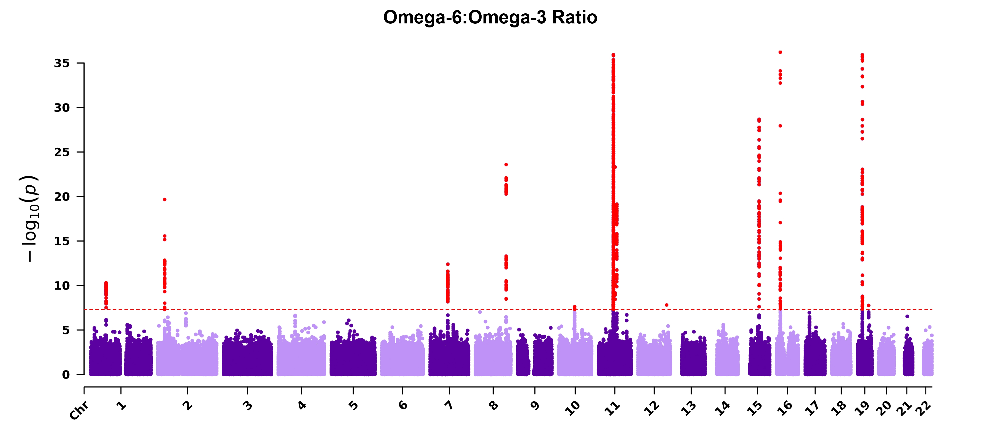

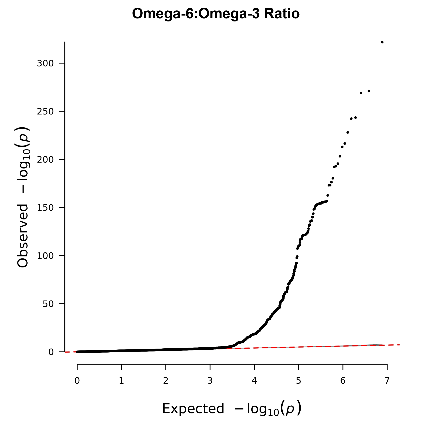

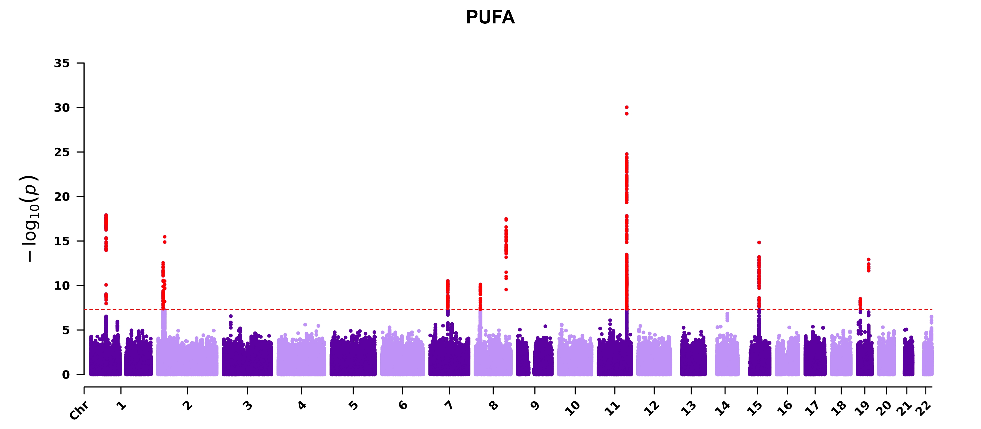

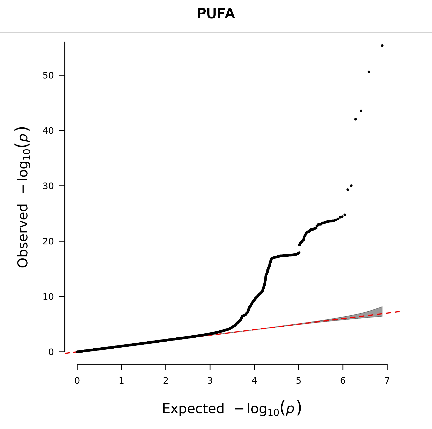

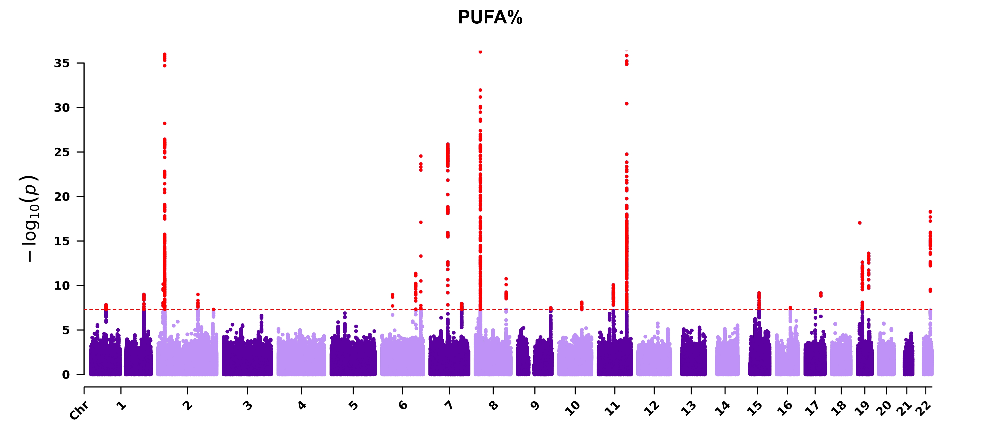

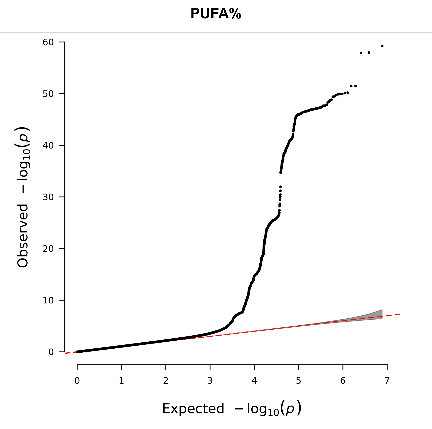

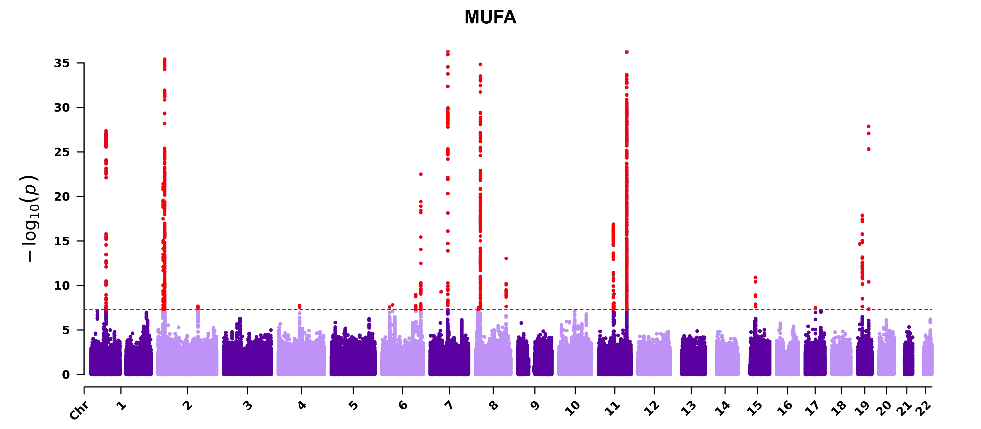

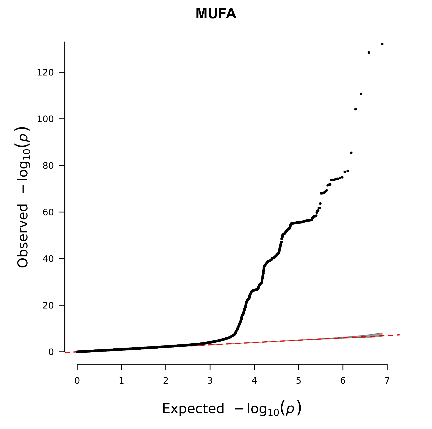

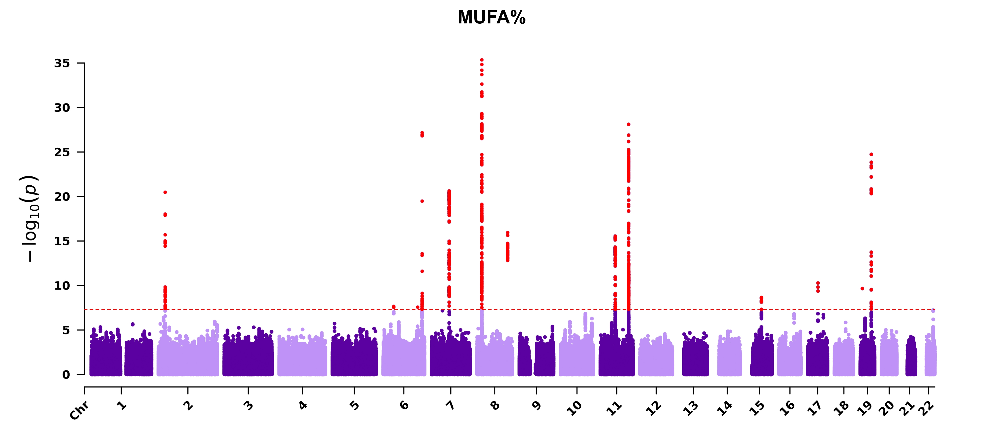

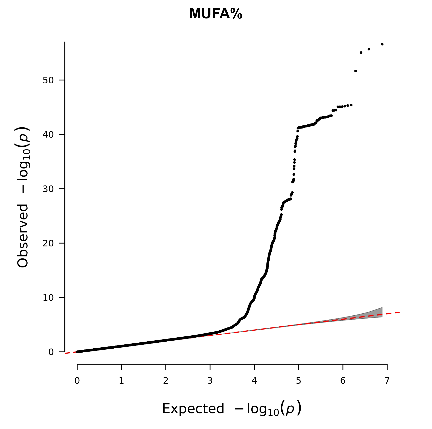

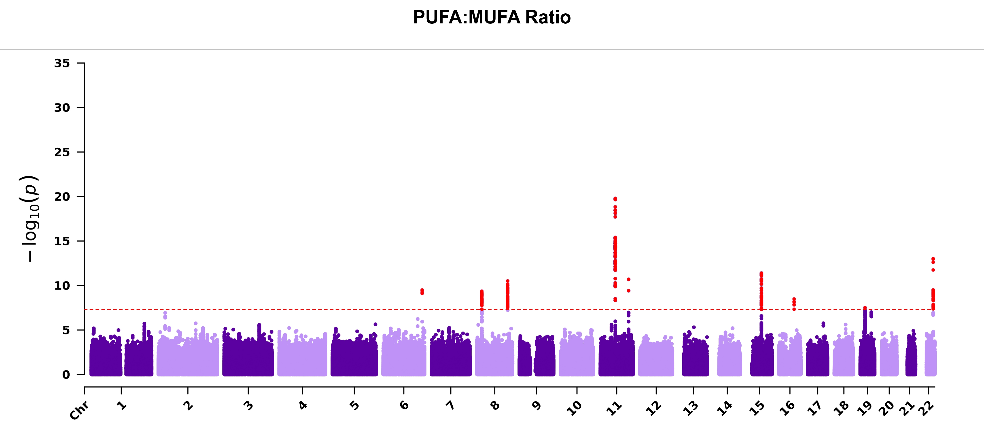

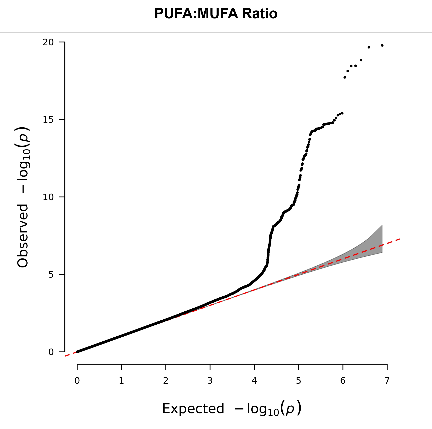

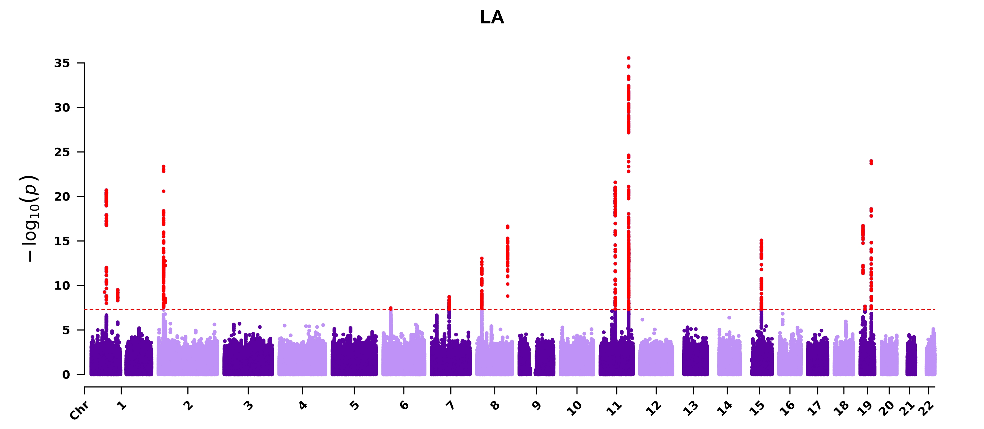

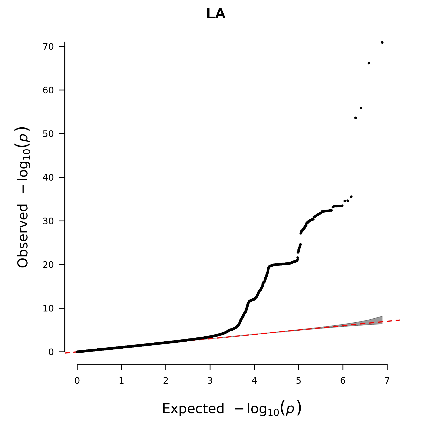

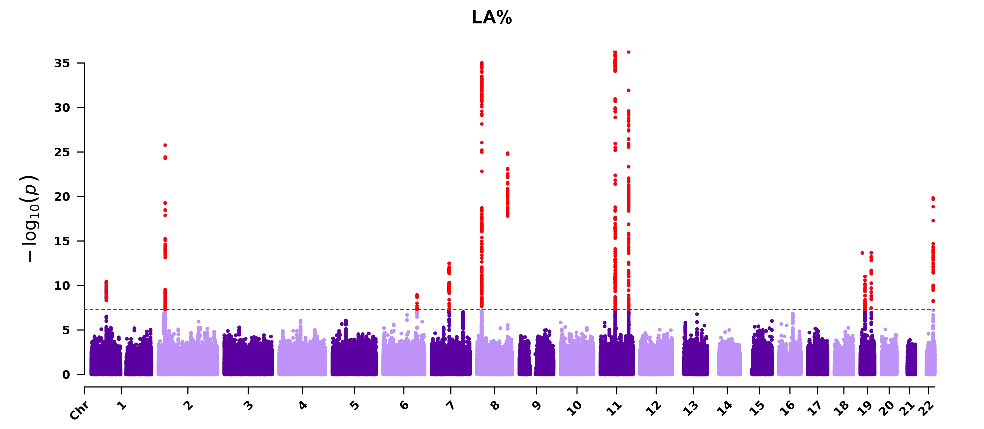

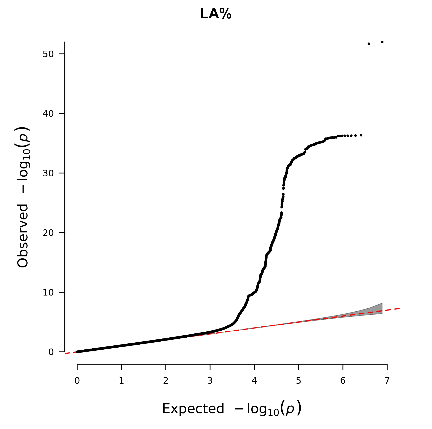

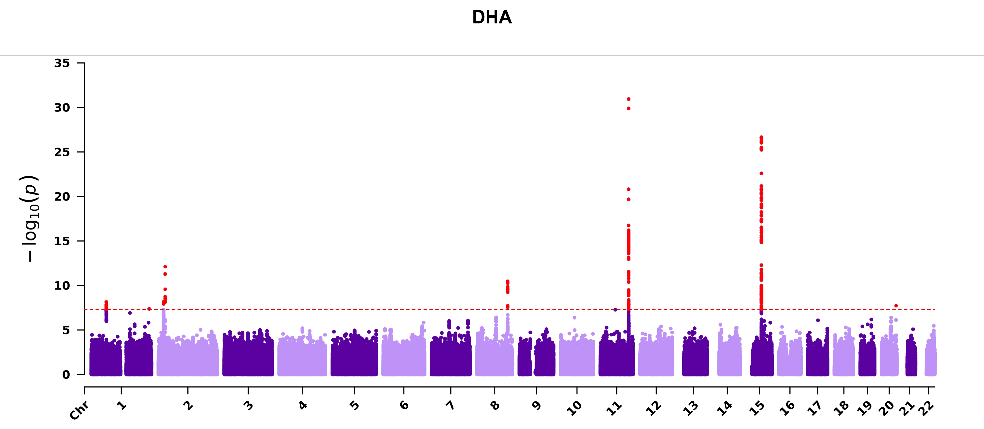

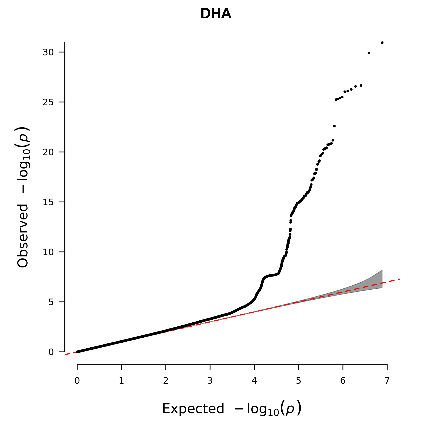

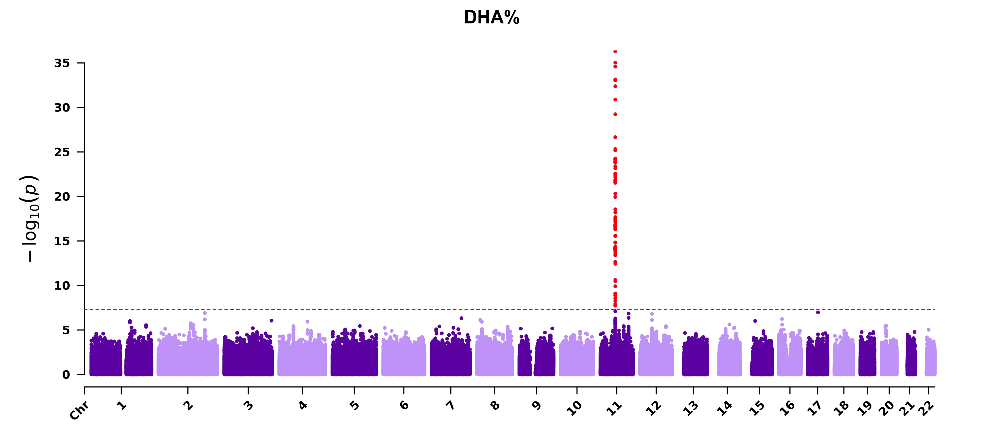

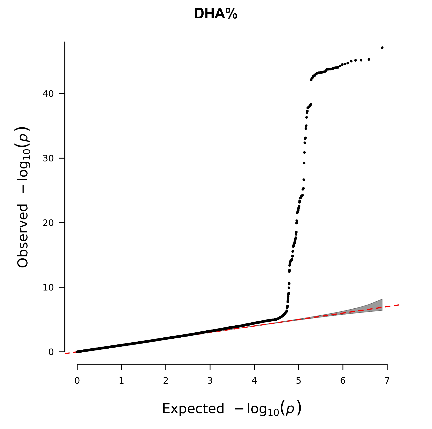

### Figure S2. Manhattan and QQ plots for vQTL analysis in 14 PUFAs- and MUFA-related traits.

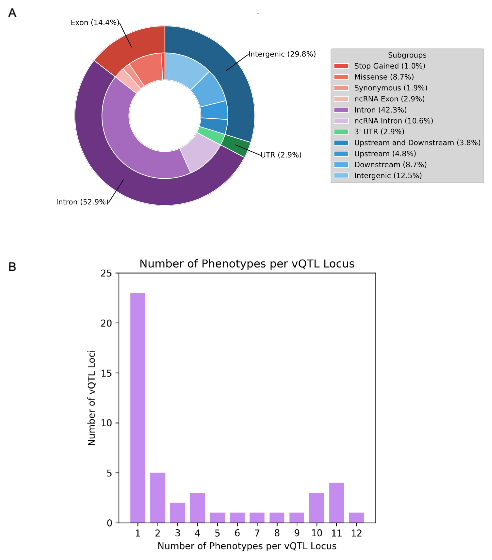

Figure S3. A) Distribution of predicted functional annotation classes for 172 independent vQTL loci. B) The number of phenotypes per non-overlapping vQTL shows that 50% of the vQTLs were significant in only one trait.

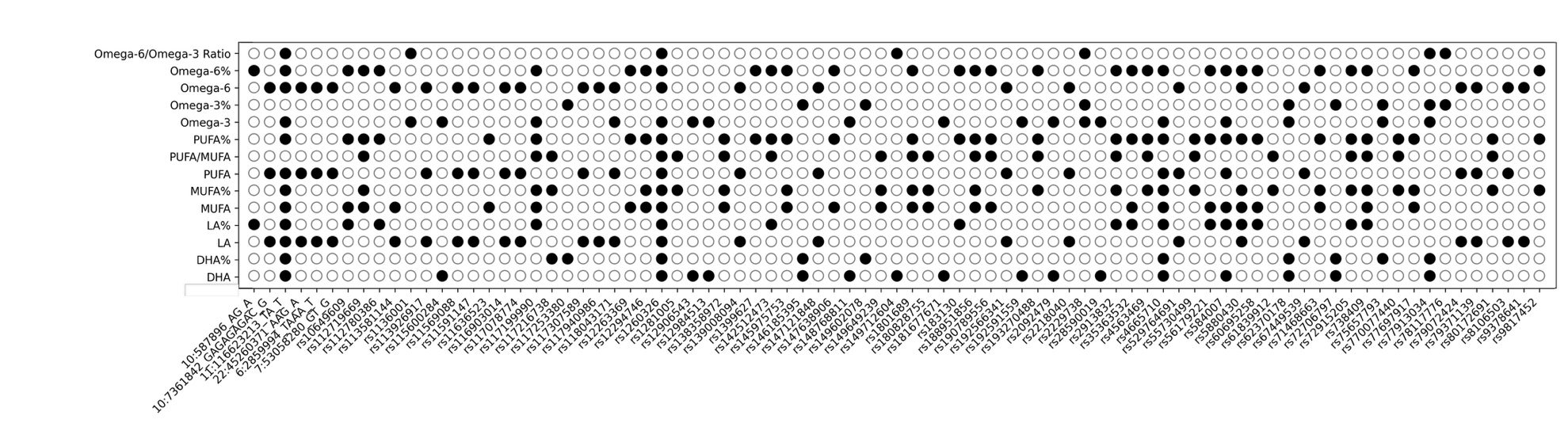

Figure S4. Dot plot matrix visualizing 83 colocalized loci across 14 PUFA- and MUFA-related traits at PP > 0.7. The total PUFAs, LA, and Omega-6 clusters shared the most loci.

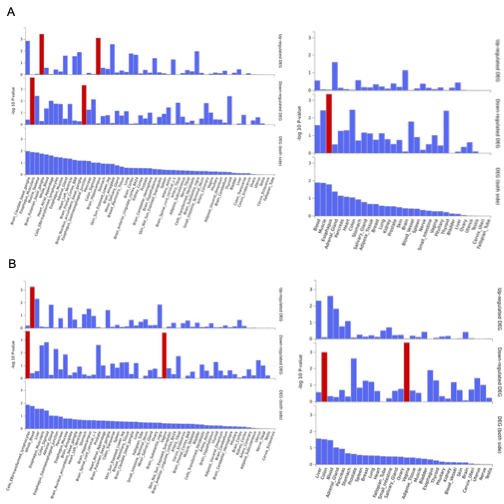

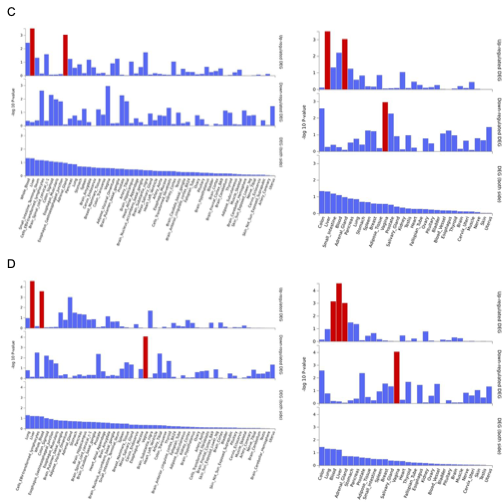

Figure S5. Tissue-specific expression of vQTL-mapped genes in A) DHA%, B) LA%, C) Omega-6%, D) PUFAs% at FDR < 0.05. Red bars indicate significant tissues. Left panels show analysis in 53 tissue types, and the right panels show analysis in 30 general tissue types. The -log10(FDR-adjusted *p*) is plotted on the y-axis

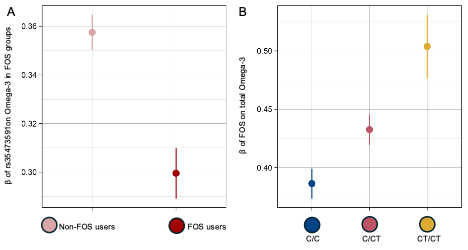

Figure S6**.** A) Marginal genetic effect of rs35473591 on Omega-3 in FOS users and non-users. B) The effect of FOS on total Omega-3 across rs35473591 genotype groups. All error bars represent 95% confidence intervals.
